# Development and Evaluation of the DMIND Questionnaire: Preparing for AI Integration into an Effective Depression Screening Tool

**DOI:** 10.1101/2024.06.07.24308625

**Authors:** Solaphat Hemrungrojn, Kittipoch Saengsai, Pasit Jakkrawankul, Chanyanart Kiattiporn-Opas, Kantapat Chaichareenon, Arisara Amrapala, Kulvara Lapanan, Sarunya Hengprapom, Narin Hiransuthikul, Titipat Achakulvisut, Natawut Nupairoj, Phanupong Phutrakool, Rapinpat Yodlorchai, Peerapol Vateekul

**Affiliations:** Center of Excellence in Digital and AI for Mental Health, Faculty of Engineering, Chulalongkorn University, Bangkok, Thailand; Cognitive Fitness and Biopsychiatry Technology Research Unit, Faculty of Medicine, Chulalongkorn University, Bangkok, Thailand; Department of Preventive and Social Medicine, Faculty of Medicine, Chulalongkorn University, Bangkok, Thailand; Department of Psychiatry, Faculty of Medicine, Chulalongkorn University, Bangkok, Thailand; Neuroscience Research Australia, Randwick, New South Wales, Australia; School of Population Health, Faculty of Medicine and Health, University of New South Wales, Sydney, New South Wales, Australia; Department of Biomedical Engineering, Faculty of Engineering, Mahidol University, Bangkok, Thailand; Department of Computer Engineering, Faculty of Engineering, Chulalongkorn University, Bangkok, Thailand; Chula Data Management Center, Faculty of Medicine, Chulalongkorn University, Bangkok, Thailand; Center of Excellence in Preventive and Integrative Medicine, Faculty of Medicine, Chulalongkorn University, Bangkok, Thailand

**Keywords:** Depression, Depression Screening, Mental Health, Artificial intelligence

## Abstract

**Objective:** Thailand’s mental health crisis is exacerbated by high demand and a shortage of mental health professionals. The research objective was to develop and validate the Detection and Monitoring Intelligence Network for Depression (DMIND) questionnaire, designed to be culturally relevant and easily administered in clinical settings. Crafted with expert input, items specifically conducive to artificial intelligence (AI) analysis were selected to facilitate the future development of an AI-assisted depression scoring model. This approach underscores the tool’s dual utility in both human-led and technology-enhanced diagnostics.

**Materials and Methods:** We enrolled 81 participants from psychiatric and tertiary care hospitals in Bangkok. Participants were assessed using the DMIND questionnaire, followed by the Hamilton Depression Rating Scale (HDRS-17). Statistical analyses included the content validity index (CVI), Cronbach’s alpha, Pearson’s correlation coefficient, Cohen’s kappa, and receiver operating characteristic (ROC) analysis. The Liu method, Youden index, and nearest neighbor method were used to determine the optimal cut-off point

**Results:** The DMIND questionnaire showed strong validity, with an item-level CVI (I-CVI) and scale-level CVI (S-CVI) exceeding 1.0, indicating strong consensus on its relevance and utility. The tool also demonstrated high internal consistency (Cronbach’s alpha = 0.96). ROC analysis showed an AUC of 0.88, indicating high accuracy in depression screening. An optimal cut-off score of 11.5 was identified, balancing predictive value and sensitivity.

**Conclusion:** The DMIND questionnaire represents a significant advancement in innovative mental health diagnostics, addressing unmet clinical needs by providing accurate and efficient assessments capable of AI integration for further enhancing mental health service delivery in Thailand.

## Introduction

Thailand is grappling with a mounting mental health crisis, characterized by a surge in depression rates that increasingly overwhelm the available healthcare resources. This scenario necessitates the rapid detection and management of depression to ensure effective intervention and mitigate risks such as suicide^1^. Unfortunately, current diagnostic practices are severely limited by the scarcity of professionals trained to employ complex tools like the Hamilton Depression Rating Scale (HDRS), the Beck Depression Inventory (BDI), and the Montgomery-Åsberg Depression Rating Scale (MADRS). These tools, while rigorous, require significant expertise and time to administer, which is unfeasible in many Thai healthcare settings^2,3,4^.

Considering these challenges, there is an urgent need to develop new, more accessible diagnostic tools. These tools should ideally align with clinical standards akin to those of the HDRS^5,6^ but with greater ease of use, capable of capturing the delicate expressions of depression. Simplicity and sensitivity are vital for artificial intelligence (AI)-driven systems, enhancing the systems’ ability to detect depression from emotional text effectively. Utilizing AI in this context expedites the diagnosis of depression and strengthens assessment precision^7,8,9^, thereby enabling quicker and more effective treatment decisions in Thai healthcare settings^10^.

Moreover, unlike the simple adaptation of existing tools, questionnaire development requires a comprehensive understanding of both the clinical landscape and AI technology^11^. This dual focus ensures that the new tool is clinically effective and optimized for AI applications, potentially revolutionizing diagnostic practices by improving accuracy and operational efficiency^12,13^ ^14^.

Taken together, the study’s objective was to construct and validate the Detection and Monitoring Intelligence Network for Depression (DMIND) questionnaire, aiming for it to be culturally relevant, easily administered in clinical settings, and seamlessly integrated with AI systems in the future.

## Materials and Methods

### Study Participants and Eligibility

Participants aged 18-65 were recruited using quota sampling from outpatient departments at two hospitals in Bangkok, Thailand: The Somdet Chaopraya Institute of Psychiatry and King Chulalongkorn Memorial Hospital. This method ensured a balanced representation of normal controls and patients diagnosed with depression. Inclusion criteria comprised proficiency in Thai, absence of intellectual disability, and consent to video and audio recording. Exclusion criteria consisted of communication difficulties, facial expression issues, or any schizophrenia spectrum disorder or substance use disorder. Data collection occurred between September and December 2023. All participants provided informed consent before data collection. Clinical depression was diagnosed by an HDRS score ≥ 8, whereas normal controls scored 7 or less.

A total of 81 participants were enrolled: 39 were diagnosed with depression, and 42 were non-depressed controls. The sample size calculation followed the method by Beam^15^, utilizing HDRS sensitivity as the gold standard (sensitivity = 0.85). The target sensitivity was 0.65, reflecting the standard sensitivity range of 0.6-0.9 commonly used in depression screening tests. The sample size was calculated to be 72 using the Sample size for comparing the sensitivity (or specificity) of two diagnostic tests .

### DMIND Questionnaire Development

The DMIND questionnaire was developed for depression screening by adapting items from the 17-item HDRS (HDRS-17)^16,17^ and the Patient Health Questionnaire-9 (PHQ-9)^18,19^. While HDRS-17 offers a comprehensive assessment of depression severity, PHQ-9 focuses on specific depression symptoms^20^. Two expert psychiatrists reviewed both scales and selected questions deemed appropriate for the study population and suitable for AI-based scoring. Two items from PHQ-9 (loss of interest, depressed mood) and four from HDRS-17 (depressed mood, loss of interest, inability to work, suicidal ideation) were selected due to their potential to evoke strong emotional responses, making them ideal for AI systems to capture nuanced linguistic and behavioral cues. These six selected items from PHQ-9 and HDRS-17 were adapted and further expanded into additional questions guiding the user/patient to provide more information and induce more emotional responses. Following pilot testing conducted in July and September 2023, the final version comprised nine items (six scored and three unscored). The DMIND questionnaire was then adapted into an audio version, integrated into an application (known as the DMIND application), and administered via an avatar resembling a psychiatrist. This digital interface enables standardized assessment and AI analysis of video responses.

### Research Procedure

Participants were interviewed at their respective hospitals, either following their doctor’s appointment or at their convenience. They first completed the DMIND questionnaire using the application, followed by assessment with the HDRS-17. Both assessments were administered by a trained psychiatric nurse/psychologist on the same day in quiet, distraction-free rooms.

Evaluators for each assessment were blinded to the results of the other. Subsequently, a licensed psychiatrist reviewed and scored the responses on the DMIND application.

### Assessments

Our DMIND questionnaire comprises nine items rated on a 4-point scale, including six scored items and three additional open-ended questions that are not scored. Higher scores indicate higher levels of depression, with a maximum value of 36 points. The following is an example questionnaire item: “คุณยังอดทนหรือฝืนทําหน้าที3หลักนัน6 ได้อยู่ไหม” (translation: “Can you still endure or resist carrying out your main duty?”). Response videos were rated by a trained psychiatrist with many years of experience.

The HDRS-17, developed by Max Hamilton in the late 1950s, is widely used in clinical trials to measure depression severity. It assesses depressive symptoms over the preceding 14 days, including depressed mood, feelings of guilt, suicide ideation, insomnia, work and activity level, retardation, agitation, psychic and somatic anxiety, gastrointestinal and general somatic symptoms, genital symptoms, hypochondriasis, weight loss, and insight. Responses are rated on a 5-point scale (0-4), with higher scores indicating greater severity. With a maximum score of 53, interpretation of scores ranges from 0-7 (normal), 8-16 (mild depression), 17-23 (moderate depression), to 24 or higher (severe depression).

### Statistical Analysis

Results were summarised using descriptive statistics. The content validity of the developed questionnaire was evaluated using the content validity index (CVI) where four independent experts graded each item for relevance and clarity. Internal consistency was evaluated using Cronbach’s alpha. The relationship between the DMIND and HDRS-17 scores was determined using Pearson’s correlation coefficient. Cohen’s kappa coefficient was used to examine the agreement between the two tools, ensuring the questionnaire’s reliability and accuracy. ROC analysis helped determine the area under the ROC curve (AUC), while the optimal cut-off point was evaluated using the Liu method^21^, Youden’s index (YI)^22^, and the nearest neighbor method. Sensitivity, specificity, positive predictive value (PPV), and negative predictive value (NPV) were also calculated. All analyses were conducted using STATA software release 15.1.

## Results

### Participant Demographics

The study included 81 participants (19 males and 62 females), comprising 42 normal controls and 39 patients with depression. The mean age was 33.91±10.53 years. Among the participants, 17.28% were married and 62.97% were employed. Most participants had completed a bachelor’s degree or higher (75.31%), whereas the rest had completed either secondary (17.28%) or elementary education (6.17%). Detailed demographic information is provided in **Table 1**.

**Table 1.**
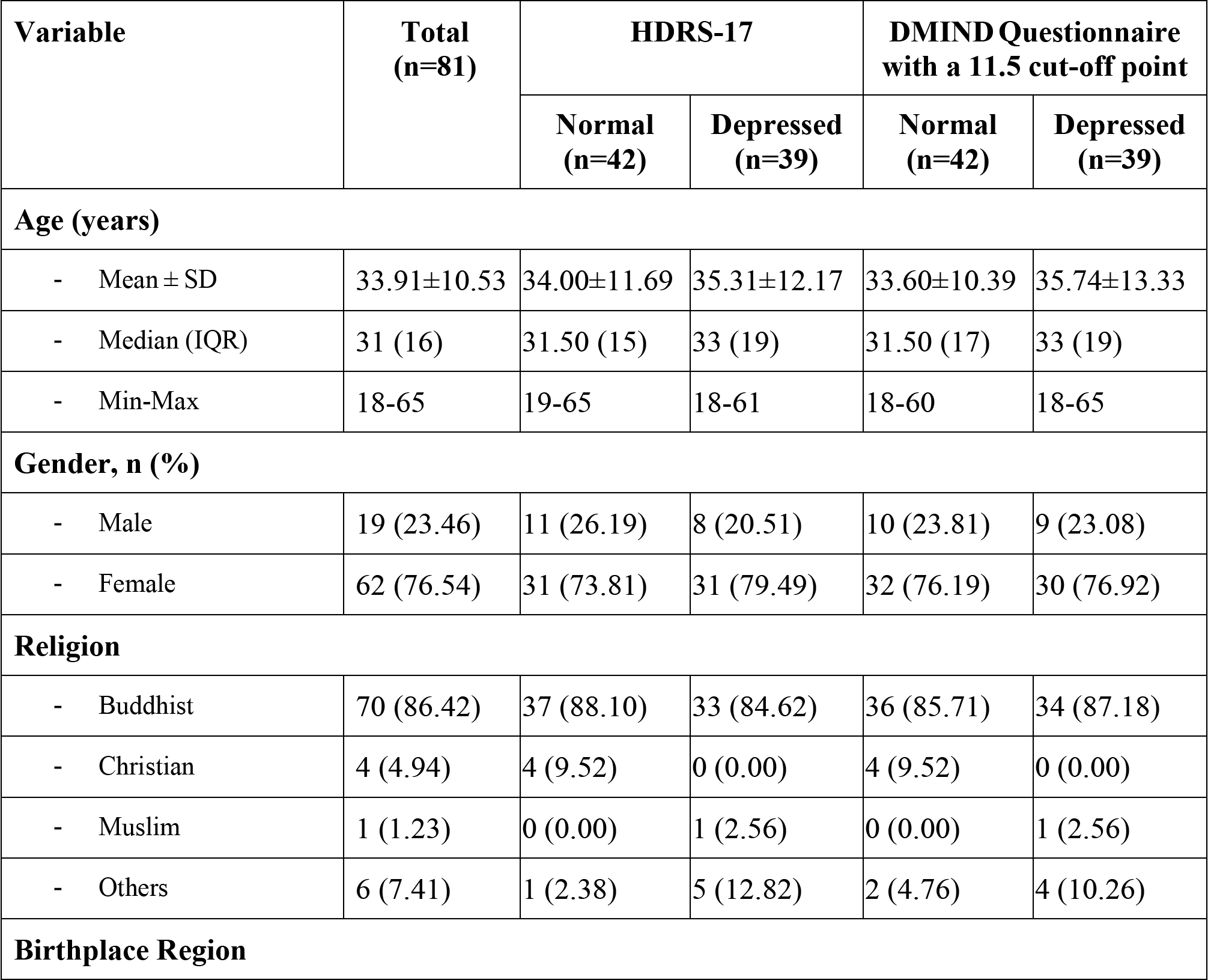
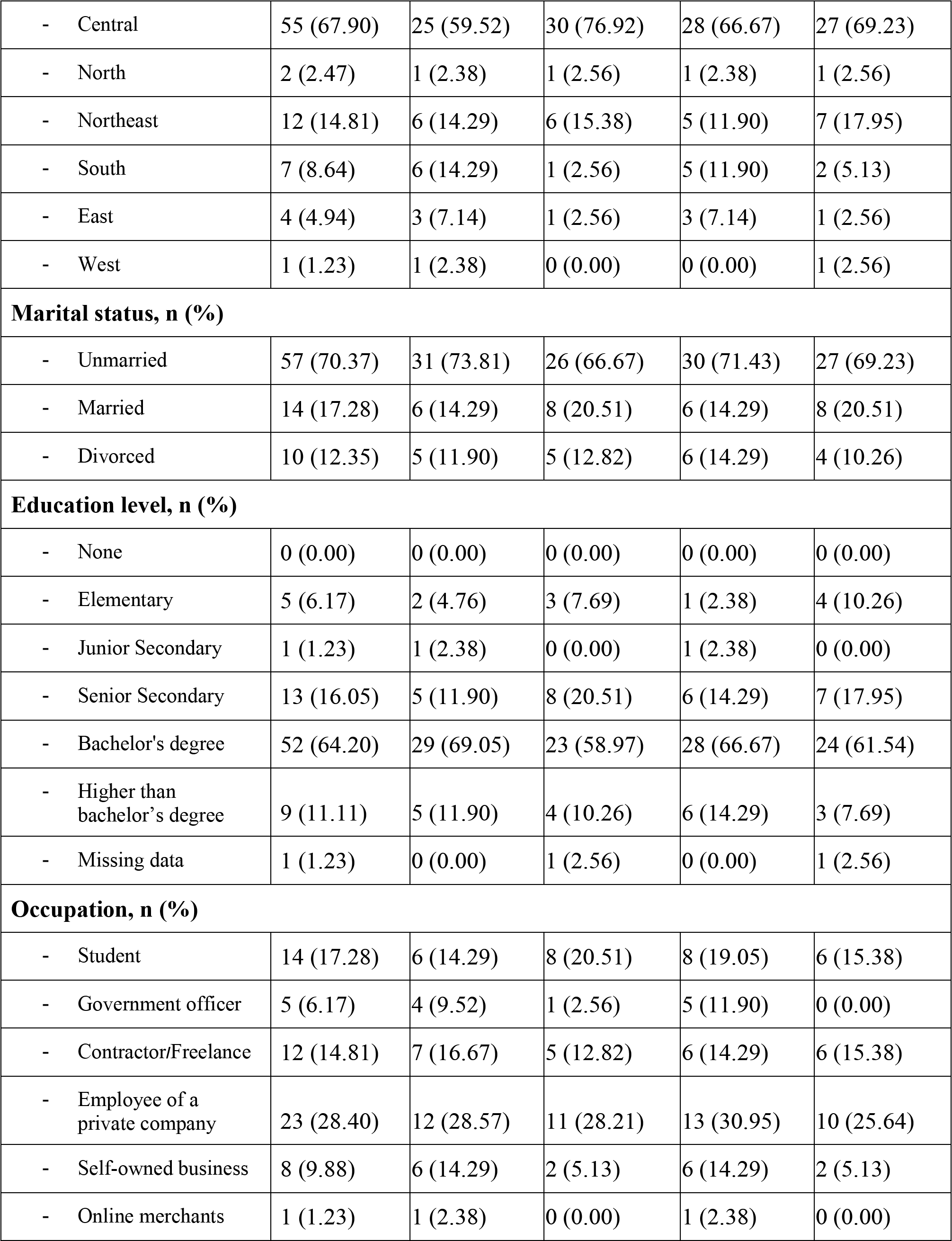
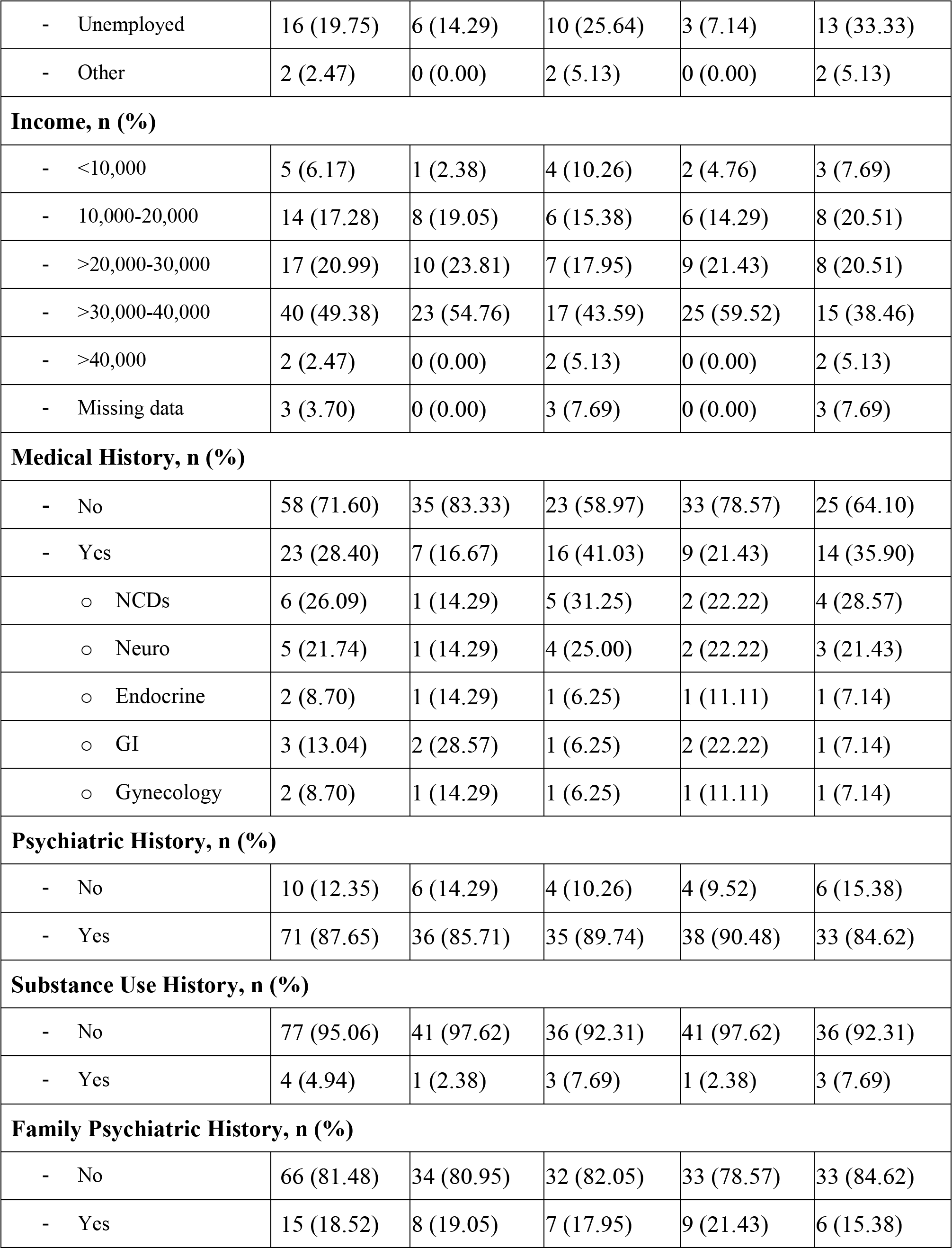
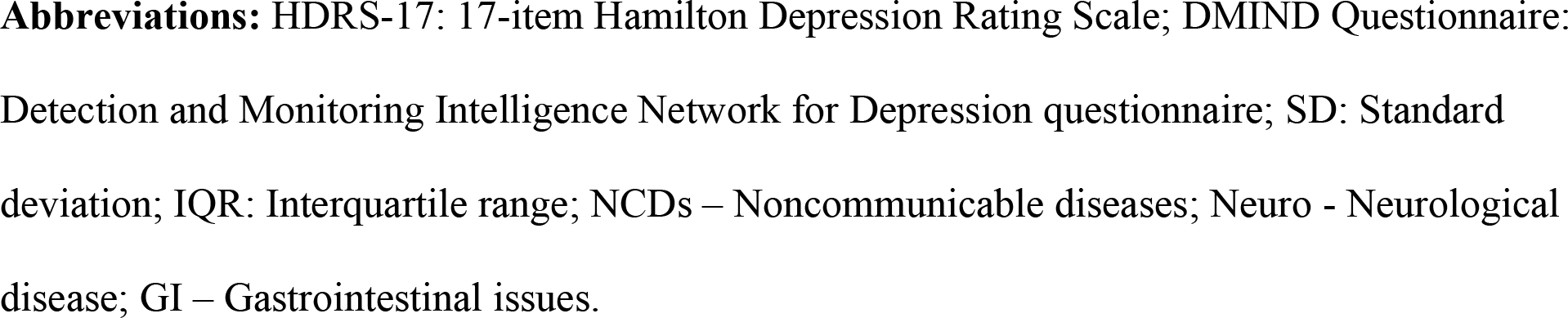
Demographic characteristics.

| Variable | Total<br>(n=81) | HDRS-17 |  | DMIND Questionnaire<br>with a 11.5 cut-off point |  |
| --- | --- | --- | --- | --- | --- |
|  |  | Normal<br>(n=42) | Depressed<br>(n=39) | Normal<br>(n=42) | Depressed<br>(n=39) |
| Age (years) |  |  |  |  |  |
| - Mean ± SD | 33.91±10.53 | 34.00±11.69 | 35.31±12.17 | 33.60±10.39 | 35.74±13.33 |
| - Median (IQR) | 31 (16) | 31.50 (15) | 33 (19) | 31.50 (17) | 33 (19) |
| - Min-Max | 18-65 | 19-65 | 18-61 | 18-60 | 18-65 |
| Gender, n (%) |  |  |  |  |  |
| - Male | 19 (23.46) | 11 (26.19) | 8 (20.51) | 10 (23.81) | 9 (23.08) |
| - Female | 62 (76.54) | 31 (73.81) | 31 (79.49) | 32 (76.19) | 30 (76.92) |
| Religion |  |  |  |  |  |
| - Buddhist | 70 (86.42) | 37 (88.10) | 33 (84.62) | 36 (85.71) | 34 (87.18) |
| - Christian | 4 (4.94) | 4 (9.52) | 0 (0.00) | 4 (9.52) | 0 (0.00) |
| - Muslim | 1 (1.23) | 0 (0.00) | 1 (2.56) | 0 (0.00) | 1 (2.56) |
| - Others | 6 (7.41) | 1 (2.38) | 5 (12.82) | 2 (4.76) | 4 (10.26) |
| Birthplace Region |  |  |  |  |  |
| - Central | 55 (67.90) | 25 (59.52) | 30 (76.92) | 28 (66.67) | 27 (69.23) |
| - North | 2 (2.47) | 1 (2.38) | 1 (2.56) | 1 (2.38) | 1 (2.56) |
| - Northeast | 12 (14.81) | 6 (14.29) | 6 (15.38) | 5 (11.90) | 7 (17.95) |
| - South | 7 (8.64) | 6 (14.29) | 1 (2.56) | 5 (11.90) | 2 (5.13) |
| - East | 4 (4.94) | 3 (7.14) | 1 (2.56) | 3 (7.14) | 1 (2.56) |
| - West | 1 (1.23) | 1 (2.38) | 0 (0.00) | 0 (0.00) | 1 (2.56) |
| **Marital status, n (%)** |  |  |  |  |  |
| - Unmarried | 57 (70.37) | 31 (73.81) | 26 (66.67) | 30 (71.43) | 27 (69.23) |
| - Married | 14 (17.28) | 6 (14.29) | 8 (20.51) | 6 (14.29) | 8 (20.51) |
| - Divorced | 10 (12.35) | 5 (11.90) | 5 (12.82) | 6 (14.29) | 4 (10.26) |
| **Education level, n (%)** |  |  |  |  |  |
| - None | 0 (0.00) | 0 (0.00) | 0 (0.00) | 0 (0.00) | 0 (0.00) |
| - Elementary | 5 (6.17) | 2 (4.76) | 3 (7.69) | 1 (2.38) | 4 (10.26) |
| - Junior Secondary | 1 (1.23) | 1 (2.38) | 0 (0.00) | 1 (2.38) | 0 (0.00) |
| - Senior Secondary | 13 (16.05) | 5 (11.90) | 8 (20.51) | 6 (14.29) | 7 (17.95) |
| - Bachelor's degree | 52 (64.20) | 29 (69.05) | 23 (58.97) | 28 (66.67) | 24 (61.54) |
| - Higher than bachelor's degree | 9 (11.11) | 5 (11.90) | 4 (10.26) | 6 (14.29) | 3 (7.69) |
| - Missing data | 1 (1.23) | 0 (0.00) | 1 (2.56) | 0 (0.00) | 1 (2.56) |
| **Occupation, n (%)** |  |  |  |  |  |
| - Student | 14 (17.28) | 6 (14.29) | 8 (20.51) | 8 (19.05) | 6 (15.38) |
| - Government officer | 5 (6.17) | 4 (9.52) | 1 (2.56) | 5 (11.90) | 0 (0.00) |
| - Contractor/Freelance | 12 (14.81) | 7 (16.67) | 5 (12.82) | 6 (14.29) | 6 (15.38) |
| - Employee of a private company | 23 (28.40) | 12 (28.57) | 11 (28.21) | 13 (30.95) | 10 (25.64) |
| - Self-owned business | 8 (9.88) | 6 (14.29) | 2 (5.13) | 6 (14.29) | 2 (5.13) |
| - Online merchants | 1 (1.23) | 1 (2.38) | 0 (0.00) | 1 (2.38) | 0 (0.00) |
| - Unemployed | 16 (19.75) | 6 (14.29) | 10 (25.64) | 3 (7.14) | 13 (33.33) |
| - Other | 2 (2.47) | 0 (0.00) | 2 (5.13) | 0 (0.00) | 2 (5.13) |
| **Income, n (%)** |  |  |  |  |  |
| - <10,000 | 5 (6.17) | 1 (2.38) | 4 (10.26) | 2 (4.76) | 3 (7.69) |
| - 10,000-20,000 | 14 (17.28) | 8 (19.05) | 6 (15.38) | 6 (14.29) | 8 (20.51) |
| - >20,000-30,000 | 17 (20.99) | 10 (23.81) | 7 (17.95) | 9 (21.43) | 8 (20.51) |
| - >30,000-40,000 | 40 (49.38) | 23 (54.76) | 17 (43.59) | 25 (59.52) | 15 (38.46) |
| - >40,000 | 2 (2.47) | 0 (0.00) | 2 (5.13) | 0 (0.00) | 2 (5.13) |
| - Missing data | 3 (3.70) | 0 (0.00) | 3 (7.69) | 0 (0.00) | 3 (7.69) |
| **Medical History, n (%)** |  |  |  |  |  |
| - No | 58 (71.60) | 35 (83.33) | 23 (58.97) | 33 (78.57) | 25 (64.10) |
| - Yes | 23 (28.40) | 7 (16.67) | 16 (41.03) | 9 (21.43) | 14 (35.90) |
| o NCDs | 6 (26.09) | 1 (14.29) | 5 (31.25) | 2 (22.22) | 4 (28.57) |
| o Neuro | 5 (21.74) | 1 (14.29) | 4 (25.00) | 2 (22.22) | 3 (21.43) |
| o Endocrine | 2 (8.70) | 1 (14.29) | 1 (6.25) | 1 (11.11) | 1 (7.14) |
| o GI | 3 (13.04) | 2 (28.57) | 1 (6.25) | 2 (22.22) | 1 (7.14) |
| o Gynecology | 2 (8.70) | 1 (14.29) | 1 (6.25) | 1 (11.11) | 1 (7.14) |
| **Psychiatric History, n (%)** |  |  |  |  |  |
| - No | 10 (12.35) | 6 (14.29) | 4 (10.26) | 4 (9.52) | 6 (15.38) |
| - Yes | 71 (87.65) | 36 (85.71) | 35 (89.74) | 38 (90.48) | 33 (84.62) |
| **Substance Use History, n (%)** |  |  |  |  |  |
| - No | 77 (95.06) | 41 (97.62) | 36 (92.31) | 41 (97.62) | 36 (92.31) |
| - Yes | 4 (4.94) | 1 (2.38) | 3 (7.69) | 1 (2.38) | 3 (7.69) |
| **Family Psychiatric History, n (%)** |  |  |  |  |  |
| - No | 66 (81.48) | 34 (80.95) | 32 (82.05) | 33 (78.57) | 33 (84.62) |
| - Yes | 15 (18.52) | 8 (19.05) | 7 (17.95) | 9 (21.43) | 6 (15.38) |

**Abbreviations:** HDRS-17: 17-item Hamilton Depression Rating Scale; DMIND Questionnaire: Detection and Monitoring Intelligence Network for Depression questionnaire; SD: Standard deviation; IQR: Interquartile range; NCDs – Noncommunicable diseases; Neuro - Neurological disease; GI – Gastrointestinal issues.

The average total HDRS-17 and DMIND score among all participants was 15.20±8.12 points and 10.28±6.12 points, respectively (**Table 2**).

**Table 2.**
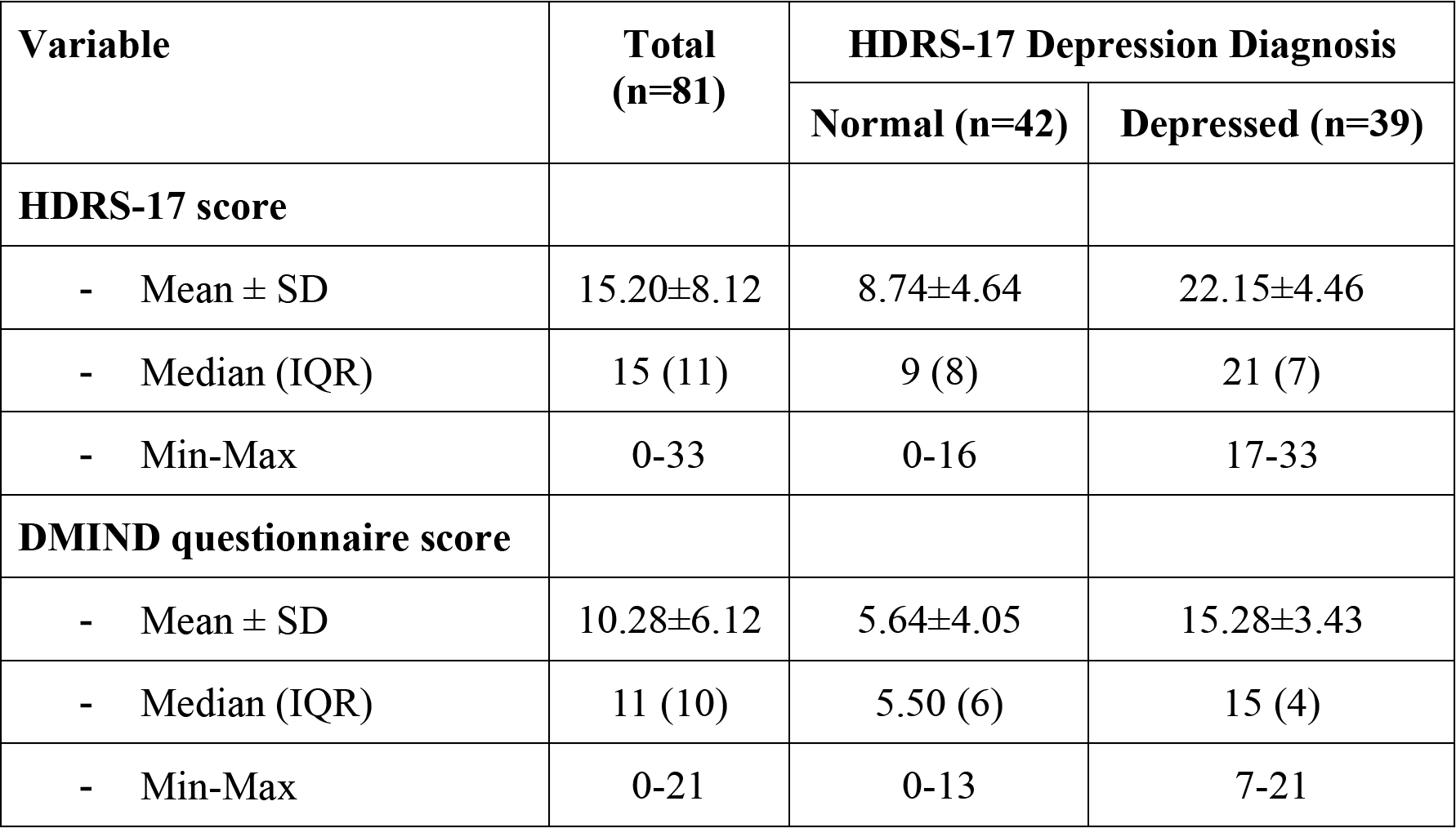
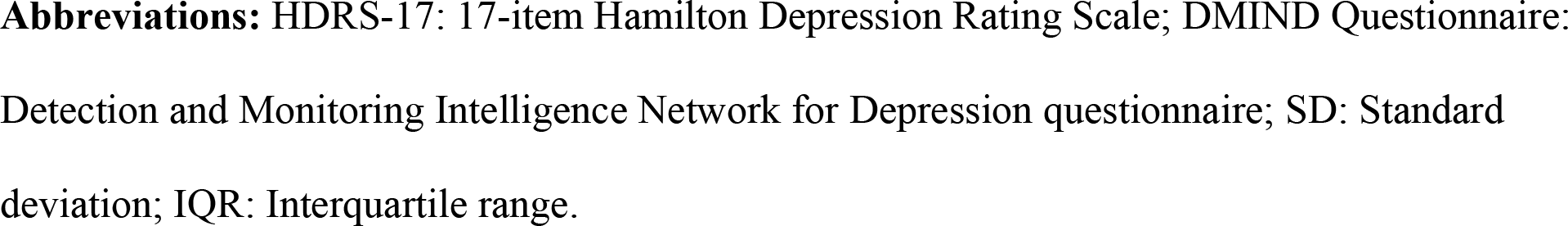
Participant scores for each assessment.

| Variable | Total<br>(n=81) | HDRS-17 Depression Diagnosis |  |
| --- | --- | --- | --- |
|  |  | Normal (n=42) | Depressed (n=39) |
| **HDRS-17 score** |  |  |  |
| - Mean ± SD | $15.20 \pm 8.12$ | $8.74 \pm 4.64$ | $22.15 \pm 4.46$ |
| - Median (IQR) | 15 (11) | 9 (8) | 21 (7) |
| - Min-Max | 0-33 | 0-16 | 17-33 |
| **DMIND questionnaire score** |  |  |  |
| - Mean ± SD | $10.28 \pm 6.12$ | $5.64 \pm 4.05$ | $15.28 \pm 3.43$ |
| - Median (IQR) | 11 (10) | 5.50 (6) | 15 (4) |
| - Min-Max | 0-21 | 0-13 | 7-21 |

**Abbreviations:** HDRS-17: 17-item Hamilton Depression Rating Scale; DMIND Questionnaire: Detection and Monitoring Intelligence Network for Depression questionnaire; SD: Standard deviation; IQR: Interquartile range.

Participants diagnosed with depression had a mean HDRS-17 score of 22.15±4.46 points and a mean DMIND score of 15.28±3.43 points. Conversely, the normal control group had a mean score of 8.74±4.64 points and 5.64±4.05 points on the HDRS-17 scale and DMIND questionnaire, respectively.

### Validity and reliability

Regarding the content validity of the DMIND questionnaire, our study found an item-level CVI (I-CVI) and a scale-level CVI (S-CVI) value of 1.0. This value indicates excellent expert consensus on each item’s relevance in measuring the intended objectives, thus demonstrating strong content validity across the questionnaire.

Regarding reliability, the DMIND questionnaire had a Cronbach’s alpha coefficient of 0.957 for the total score (see **Table 3**). The high coefficient value represents acceptable internal consistency, implying that the items within the questionnaire are sufficiently correlated and provide a consistent measure of the construct it is intended to assess.

**Table 3.**
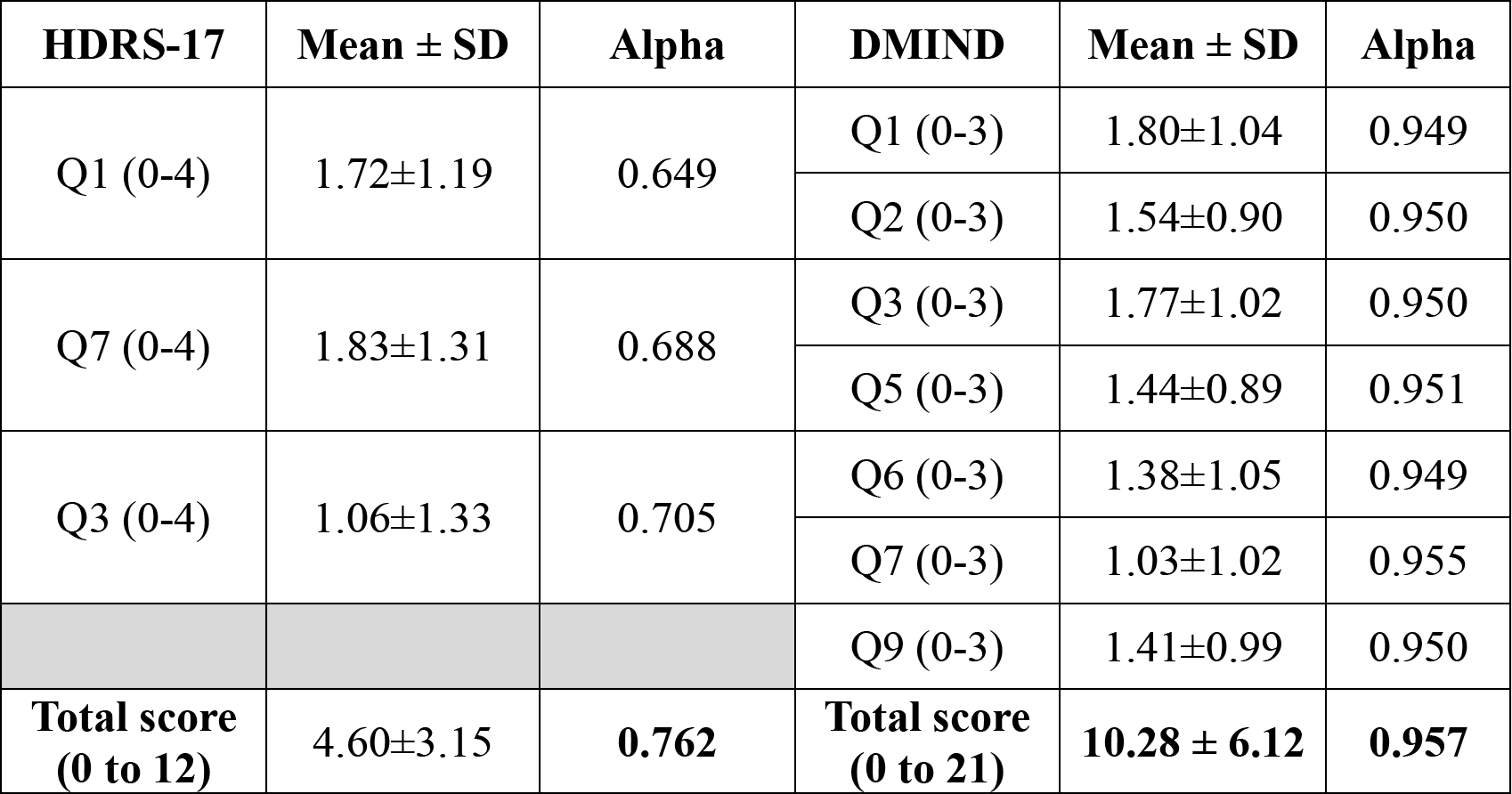
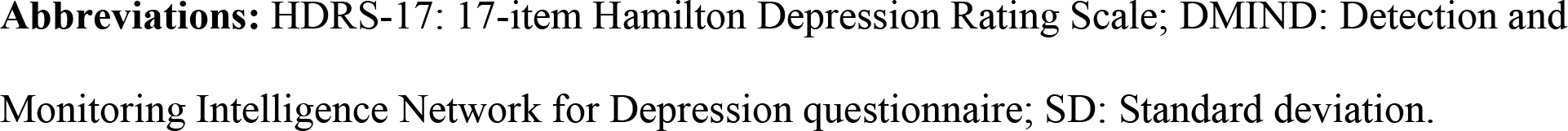
The internal consistency of the HDRS-17 and DMIND.

| **HDRS-17** | **Mean ± SD** | **Alpha** | **DMIND** | **Mean ± SD** | **Alpha** |
| --- | --- | --- | --- | --- | --- |
| Q1 (0-4) | 1.72±1.19 | 0.649 | Q1 (0-3) | 1.80±1.04 | 0.949 |
|  |  |  | Q2 (0-3) | 1.54±0.90 | 0.950 |
| Q7 (0-4) | 1.83±1.31 | 0.688 | Q3 (0-3) | 1.77±1.02 | 0.950 |
|  |  |  | Q5 (0-3) | 1.44±0.89 | 0.951 |
| Q3 (0-4) | 1.06±1.33 | 0.705 | Q6 (0-3) | 1.38±1.05 | 0.949 |
|  |  |  | Q7 (0-3) | 1.03±1.02 | 0.955 |
|  |  |  | Q9 (0-3) | 1.41±0.99 | 0.950 |
| **Total score**<br>**(0 to 12)** | **4.60±3.15** | **0.762** | **Total score**<br>**(0 to 21)** | **10.28 ± 6.12** | **0.957** |

**Abbreviations:** HDRS-17: 17-item Hamilton Depression Rating Scale; DMIND: Detection and Monitoring Intelligence Network for Depression questionnaire; SD: Standard deviation.

**Table 4.**
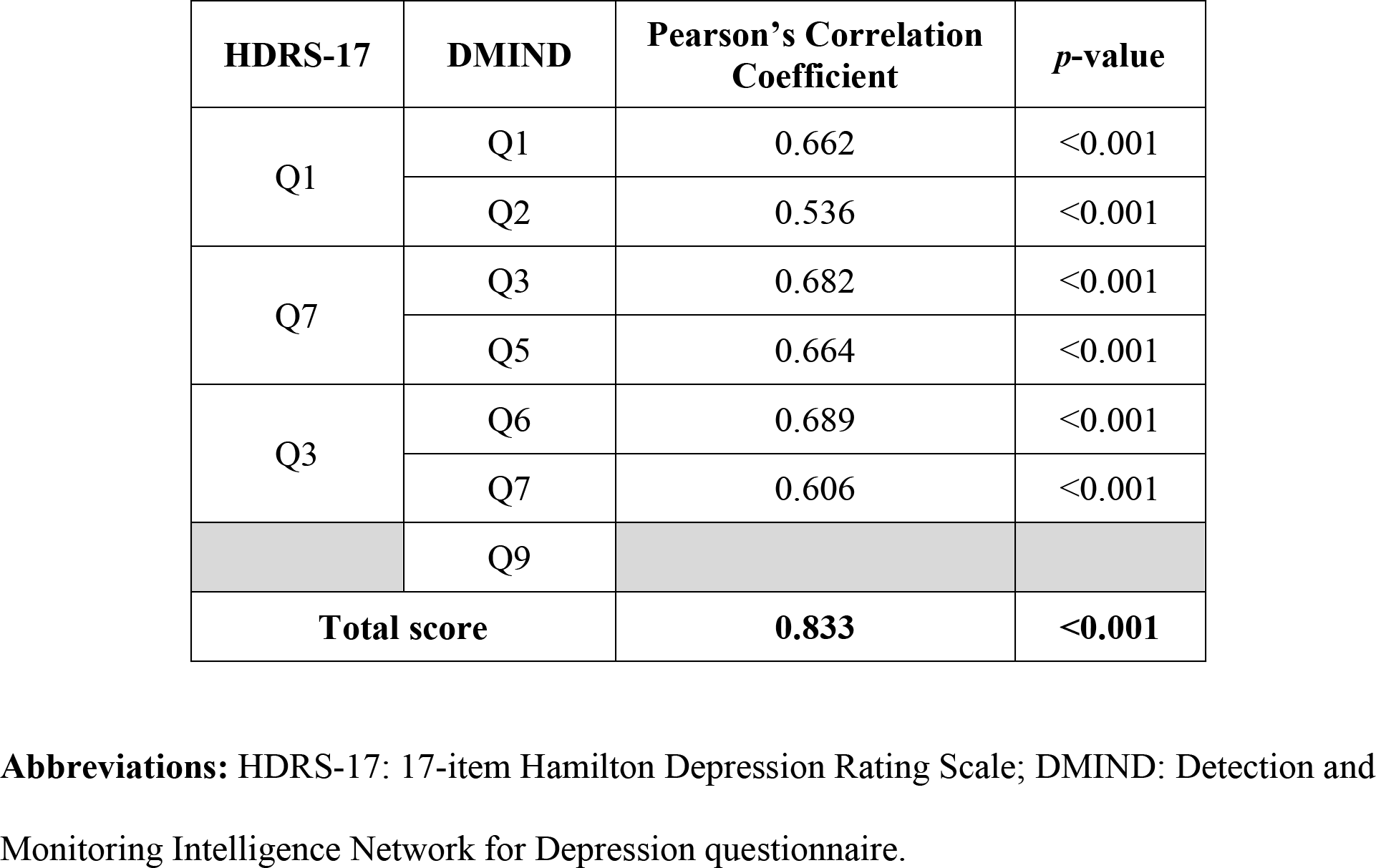
Correlation Between DMIND and HDRS-17 Total Scores.

| HDRS-17 | DMIND | Pearson's Correlation Coefficient | *p*-value |
| --- | --- | --- | --- |
| Q1 | Q1 | 0.662 | <0.001 |
|  | Q2 | 0.536 | <0.001 |
| Q7 | Q3 | 0.682 | <0.001 |
|  | Q5 | 0.664 | <0.001 |
| Q3 | Q6 | 0.689 | <0.001 |
|  | Q7 | 0.606 | <0.001 |
|  | Q9 |  |  |
| **Total score** |  | **0.833** | **<0.001** |

**Abbreviations:** HDRS-17: 17-item Hamilton Depression Rating Scale; DMIND: Detection and Monitoring Intelligence Network for Depression questionnaire.

The Pearson’s correlation coefficient between the DMIND and HDRS-17 total score was 0.833 (*p* < 0.001), signifying a strong positive correlation between the two measures. Furthermore, each item in the DMIND questionnaire had a statistically significantly high correlation with its respective HDRS-17 item. These findings support the concurrent validity of the DMIND questionnaire.

### ROC analysis and optimal cut-off point

From the ROC analysis and the nearest neighbor method, the optimal cut-off point was 11.5 points, with an AUC of 0.88 (95% CI = 0.80-0.95). At this optimal value, the sensitivity was 87.2% (95% CI = 72.6%-95.7%), the specificity was 88.1% (95% CI = 74.4%-96.0%), the PPV was 87.2% (95% CI = 72.6%-95.7%), and the NPV was 88.1% (95% CI = 74.4%-96.0%) (**Table 5**). This AUC value indicates that the DMIND questionnaire performs well in identifying patients with depression, demonstrating high sensitivity (**Figure 1**).

**Figure 1.**
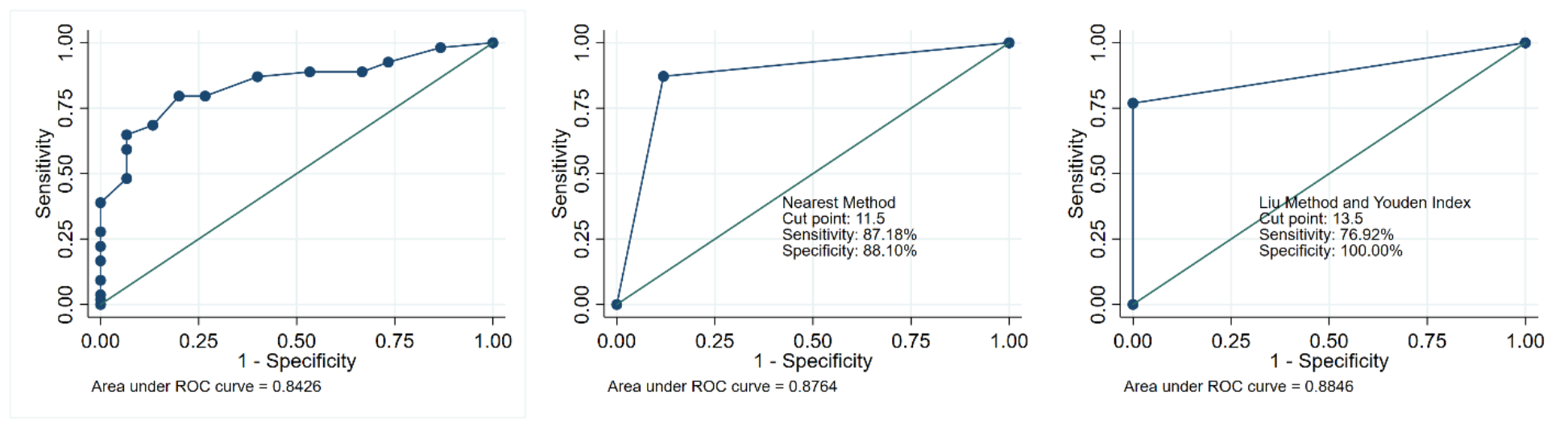
Receiver operating characteristic (ROC) analysis and optimal cut-off point using the nearest neighbor method, Liu method, and Youden index

**Table 5.** Analysis of different cut-off points using the ROC analysis.

| **Cut point** | **Sensitivity** | **Specificity** | **Correctly Classified** | **LR+** | **LR-** |
| --- | --- | --- | --- | --- | --- |
| ≥ 0 | 100.00% | 0.00% | 48.15% | 1.00 |  |
| ≥ 1 | 100.00% | 16.67% | 56.79% | 1.20 | 0.00 |
| ≥ 2 | 100.00% | 19.05% | 58.02% | 1.24 | 0.00 |
| ≥ 3 | 100.00% | 26.19% | 61.73% | 1.35 | 0.00 |
| ≥ 4 | 100.00% | 30.95% | 64.20% | 1.45 | 0.00 |
| ≥ 5 | 100.00% | 42.86% | 70.37% | 1.75 | 0.00 |
| ≥ 6 | 100.00% | 50.00% | 74.07% | 2.00 | 0.00 |
| ≥ 7 | 100.00% | 57.14% | 77.78% | 2.33 | 0.00 |
| ≥ 8 | 97.44% | 69.05% | 82.72% | 3.15 | 0.04 |
| ≥ 9 | 94.87% | 78.57% | 86.42% | 4.43 | 0.07 |
| ≥ 10 | 94.87% | 80.95% | 87.65% | 4.98 | 0.06 |
| ≥ 11 | 89.74% | 80.95% | 85.19% | 4.71 | 0.13 |
| $\geq 12^*$ | 87.18% | 88.10% | 87.65% | 7.32 | 0.15 |
| $\geq 13$ | 76.92% | 95.24% | 86.42% | 16.15 | 0.24 |
| $\geq 14^{**}$ | 76.92% | 100.00% | 88.89% | | 0.23 |
| $\geq 15$ | 64.10% | 100.00% | 82.72% | | 0.36 |
| $\geq 16$ | 48.72% | 100.00% | 75.31% | | 0.51 |
| $\geq 17$ | 30.77% | 100.00% | 66.67% | | 0.69 |
| $\geq 18$ | 28.21% | 100.00% | 65.43% | | 0.72 |
| $\geq 19$ | 23.08% | 100.00% | 62.96% | | 0.77 |
| $\geq 20$ | 12.82% | 100.00% | 58.02% | | 0.87 |
| $\geq 21$ | 2.56% | 100.00% | 53.09% | | 0.97 |
| $> 21$ | 0.00% | 100.00% | 51.85% | | 1 |

\* Optimal cut-off point using the nearest neighbor method was 11.5 points
\*\* Optimal cut-off point using the Liu method and Youden index was 13.5 points
**Abbreviations:** ROC: Receiver operating characteristic; LR+: Positive likelihood ratios; LR-: Negative likelihood ratios.

Using the Liu method and Youden index, the optimal cut-off score was 13.5 points with an AUC of 0.88 (95% CI = 0.82-0.95). At this value, the sensitivity was 76.9% (95% CI = 60.7%-88.9%), the specificity was 100% (95% CI = 91.6%-100%), the PPV was 100% (95% CI = 88.4%-100%), and the NPV was 82.4% (95% CI = 69.1%-91.6%).

### Agreement Between the DMIND questionnaire and the HDRS-17

The Cohen’s kappa statistic was calculated for both optimal cut-off scores determined in the previous analysis. When the cut-off score was 11.5 points, the percentage agreement was 87.65 percent and the Kappa value (κ) was 0.75, suggesting substantial agreement (*<u>p</u>* < 0.001) between the two tools. Similarly, for a cut-off score of 13.5 points, the percentage agreement was 88.89 percent and κ = 0.78, indicating substantial agreement (*p* < 0.001) between the tools.

These findings further support the reliability of the DMIND questionnaire, signifying strong alignment between the DMIND questionnaire and HDRS-17 categories.

## Discussion

### The DMIND questionnaire is a valid tool for depression pre-screening

The DMIND questionnaire demonstrates high reliability for depression pre-screening, as indicated by a Cronbach’s alpha coefficient of 0.957, signifying strong internal consistency. This reliability is comparable to other assessments, such as the Thai HDRS-17^16^. Key items like depressed mood and suicidal thoughts in the DMIND questionnaire were intentionally developed to yield higher scores, aiming to ensure timely intervention and minimize false negatives. Furthermore, the ROC curve analysis revealed a promising AUC of 0.88, placing our tool in a favorable position among existing tools like the MADRS, which has an AUC of 0.78^23,24,25,26,27,28^. These findings indicate that the DMIND questionnaire effectively distinguishes between depressed and non-depressed individuals.

An optimal cut-off score of 11.5 points resulted in a sensitivity of 87.2% and specificity of 88.1%, effectively balancing true positives and negatives. Although another optimal cut-off point was identified at 13.5 points, we selected the 11.5 cut-off score to prioritize high sensitivity, which ensures suitability for future AI integration and prevents urgent cases from being overlooked.

Looking at existing depression detection models, a study by Mudasir^29^ proposed an advanced deep learning model utilizing Word2Vec (a technique in natural language processing) and term frequency-inverse document frequency (TF-IDF). These methods were employed to train convolutional neural network (CNN) and long short-term memory (LSTM) models for early depression detection, targeting sensitive cues indicative of serious issues like self-harm or suicidal thoughts, which current depression detection models often fail to accurately detect. The study collected data from Facebook, Twitter, and YouTube using advanced crawling strategies to ensure a unique, diverse dataset with various indicators of depression. The authors reported that the Word2Vec LSTM and Word2Vec (CNN + LSTM) models achieved accuracies of 99.02% and 99.01%, respectively, outperforming existing methods in recall, precision, accuracy, and F1-score. Word2Vec features were particularly effective, achieving accuracies of 95.02% (CNN) and 98.15% (CNN + LSTM) on Facebook and YouTube data. In another study, sensitivity analysis results from Guohou, Lina, and Dongsong^30^ revealed that problem-related questions were the most influential in depression detection. More specifically, questions about depression, emotions, and unresolved life problems significantly impacted detection accuracy when multimodal features were utilized. Additionally, Amanat et al.^31^ proposed a productive model by implementing the LSTM and recurrent neural network (RNN) model to predict depression from text, semantics, and written content derived from interviews with 99.0% accuracy, thereby proving beneficial in protecting suicidal individuals. Together, these approaches support our current perspective of the DMIND questionnaire, where we hope to collect detailed video responses and apply AI techniques to optimize depression detection

### Practical uses of the DMIND questionnaire for future AI application

The DMIND questionnaire presents a promising tool for routine depression screening across various healthcare settings and online channels. Avatar-based interviews within our DMIND application eliminate the necessity for human involvement, reducing the workload on healthcare personnel and helping users/patients feel more comfortable expressing their true feelings and opinions. This interface enables healthcare providers to gain more in-depth information and more accurately diagnose patients. With future AI integration, the DMIND questionnaire and application have the potential to enhance depression detection from emotional text, boosting diagnostic precision and expediting treatment decisions. This fusion of clinical insight and AI technology optimizes diagnostic practices, transforming and streamlining healthcare delivery.

### Strengths and weaknesses

The DMIND questionnaire boasts several notable strengths. Questionnaire items were selected from well-established tools, and the assessor of each tool in this study was blinded to the results of the other assessment. The questionnaire demonstrated high internal consistency and strong diagnostic performance, showing high sensitivity (87.2%) and specificity (88.1%). Additionally, the questionnaire’s user-friendly design, featuring an audio interface and psychiatrist avatar, increases user engagement and comfort, potentially improving the accuracy and detail of responses compared to conventional methods. Furthermore, its potential for AI integration allows for future automated emotional text analysis, improving diagnostic precision and efficiency.

The study’s limitations include a relatively small sample size of 81 participants recruited from only two locations. Consequently, our research findings may not be generalizable to the broader Thai population. Potential bias from relying on expert review for validation could affect objectivity, highlighting the need for additional validation methods. Moreover, the questionnaire focused on a limited number of high-impact questions, which could cause other relevant symptoms to be overlooked, potentially reducing the assessment’s comprehensiveness.

### Future Research

Future research should aim to develop and integrate an AI-assisted scoring model to confirm the questionnaire’s suitability for AI integration and verify that the use of AI will effectively improve depression diagnosis precision and efficiency. Additionally, further research should be conducted to enhance the DMIND application to increase user acceptance and user-friendliness. Large-scale testing with a more diverse pool of participants is essential to validate these improvements.

## Conclusions

In conclusion, the authors successfully developed the DMIND questionnaire, a 9-item depression pre-screening tool. The developed tool exhibits strong internal consistency, discriminatory ability, and practical attributes, making it reliable for depression screening and future AI integration. Further studies will focus on developing and integrating an AI depression scoring model.

## Data Availability

All data produced in the present work are contained in the manuscript

## Acknowledgments

Firstly, we would like to thank Assoc. Prof. Chanchai Sittipun, Dean of the Faculty of Medicine, Chulalongkorn University, for his invaluable support and guidance. Secondly, we would like to express our gratitude to Dr. Manote Lotrakul, M.D. for allowing us to use the Thai version of the HDRS-17. Thirdly, we extend our thanks to the engineering and data scientist team from the Center of Excellence in Digital and AI for Mental Health (AIMET), Faculty of Engineering, Chulalongkorn University, for their continuous support in the development and implementation of our application. We are also grateful to the nurses, research assistants, and staff at King Chulalongkorn Memorial Hospital and Somdet Chaopraya Institute of Psychiatry for their cooperation and assistance. Lastly, we would like to express our deepest gratitude to the patients for participating in this study.

## Ethics Statement

All subjects gave their informed consent before participating in the study. This study was conducted in accordance with the World Medical Association Declaration of Helsinki and was approved by the Ethical Review Board of the Somdet Chaopraya Institute of Psychiatry (008/2566) and the Faculty of Medicine Chulalongkorn University (COA No. 1266/2023).

## Author Contributions

SH was responsible for the conceptualization and study design, as well as the initial drafting of the manuscript. KL contributed to the data collection and curation. PP performed the statistical analysis. KS, CK, KC, and PJ assisted in project administration. PV, TA, and NN provided supervision and guidance throughout the project. SH, NH, AA, and RY reviewed and edited the manuscript. All authors participated in the methodology design, contributed to the data interpretation, and approved the final manuscript.

## References

1. Gotlib IH, Hammen CL. Handbook of depression. 2002.Isacsson G. Suicide prevention--a medical breakthrough? Acta Psychiatr Scand. 2000;102(2):113-7. doi:10.1034/j.1600-0447.2000.102002113.x.

2. Nochaiwong S, Ruengorn C, Thavorn K,et al. Global prevalence of mental health issues among the general population during the coronavirus disease-2019 pandemic: a systematic review and meta-analysis. Sci Rep. 2021;11(1):10173. doi:10.1038/s41598-021-89700-8.

3. Prasartpornsirichoke J, Pityaratstian N, Poolvoralaks C, et al. The prevalence and economic burden of treatment-resistant depression in Thailand. BMC Public Health. 2023;23(1):1541. doi:10.1186/s12889-023-16477-y

4 . Mitchell AJ, Vaze A, Rao S. Clinical diagnosis of depression in primary care: a meta-analysis. Lancet. 2009;374(9690):609-19. doi:10.1016/s0140-6736(09)60879-5.

5. Carrozzino D, Patierno C, Fava GA, et al. The Hamilton Rating Scales for Depression: A Critical Review of Clinimetric Properties of Different Versions. Psychother Psychosom. 2020;89(3):133–50. doi:10.1159/000506879

6 Mathers CD, Loncar D. Projections of global mortality and burden of disease from 2002 to 2030. PLoS Med.2006; 3(11): e442. 10.1371/journal.pmed.0030442

6 Coverage T, Mental Health Hotline 1323.2003 https://www.thecoverage.info/news/content/4691

8 Dibeklioglu H, Hammal Z, Cohn JF. Dynamic Multimodal Measurement of Depression Severity Using Deep Autoencoding. IEEE J Biomed Health Inform.2018; 22(2):525–536. 10.1109/jbhi.2017.2676878

9 Jiang Z, Seyedi S, Griner E, et al. Multimodal Mental Health Digital Biomarker Analysis from Remote Interviews using Facial, Vocal, Linguistic, and Cardiovascular Patterns. IEEE J Biomed Health Inform.2024;Pp. 10.1109/jbhi.2024.3352075

10 Lee EE, Torous J, De Choudhury M, et al. Artificial Intelligence for Mental Health Care: Clinical Applications, Barriers, Facilitators, and Artificial Wisdom. Biol Psychiatry Cogn Neurosci Neuroimaging.2021; 6(9), 856–864. 10.1016/j.bpsc.2021.02.001

11 Williams SZ, Chung GS, Muennig PA. Undiagnosed depression: A community diagnosis. SSM Popul Health.2017;3.633–638. 10.1016/j.ssmph.2017.07.012

12 Rajpurkar P, Chen E, Banerjee O., et al. AI in health and medicine. Nat Med. 2022; 28(1), 31–38. 10.1038/s41591-021-01614-0

13. Shin D, Cho WI, Park CHK, et al. Detection of Minor and Major Depression through Voice as a Biomarker Using Machine Learning. J Clin Med.2021;10(14). 10.3390/jcm10143046

14 .Spitzer RL, Kroenke K, Williams JB. Validation and utility of a self-report version of PRIME-MD: the PHQ primary care study. Primary Care Evaluation of Mental Disorders. Patient Health Questionnaire. JAMA.1999;282(18), 1737–1744. 10.1001/jama.282.18.1737

15. Beam CA. Strategies for improving power indiagnostic radiology research.AmericanJournal of Roentgenology.1992;159: 631–37.

16. Hamilton M. A rating scale for depression. Journal of Neurology. Neurosurgery & Psychiatry. 1960;23, 56–61. 10.1136/jnnp.23.1.56

17. Lotrakul M, Sukanich P, Sukying C. The Reliability and Validity of Thai version of Hamilton Rating Scale for Depression. Journal of the Psychiatrist Association of Thailand.1996; 41(4), 235–246.

18. Kroenke K, Spitzer RL, Williams JB. The PHQ-9: validity of a brief depression severity measure. J Gen Intern Med. 2001; 16(9), 606–613. 10.1046/j.1525-1497.2001.016009606.x

19. Sumrithe S, Saipanish R. Reliability and validity of the Thai version of the PHQ-9. BMC Psychiatry.2008; 8, 46. 10.1186/1471-244x-8-46

20 .Gilbody S, Richards D, Brealey S, et al. Screening for depression in medical settings with the Patient Health Questionnaire (PHQ): a diagnostic meta-analysis. J Gen Intern Med.2007.

21. Liu X. Classification accuracy and cut point selection. Statistics in medicine. 2012;31(23), 2676– 2686. 10.1002/sim.4509

22. Youden WJ. Index for rating diagnostic tests. Cancer. 1950;3(1), 32–35. 10.1002/1097-0142(1950)3:1%3C32::AID-CNCR2820030106%3E3.0.CO;2-3

23. Beck AT, Ward CH, Mendelson M, et al. An inventory for measuring depression. Archives of General Psychiatry.1961; 4(6), 561–571. 10.1001/archpsyc.1961.01710120031004

24. Radloff LS. The CES-D Scale: A self-report report depression scale for research in the general population. Applied Psychological Measurement. 1977;1(3), 385–401. 10.1177/014662167700100306

25. Montgomery SA, Åsberg M. A new depression scale designed to be sensitive to change. The British Journal of Psychiatry.1979;134, 382–389. 10.1192/bjp.134.4.382

26. Torres SC, Olofsdotter S, Vadlin S, Ramklint M, Nilsson KW, & Sonnby K. Diagnostic accuracy of the Montgomery–Åsberg Depression Rating Scale parent report among adolescent psychiatric outpatients. Nordic Journal of Psychiatry.2017; 72(3), 184–190. 10.1080/08039488.2017.1414873

27. Spitzer RL, Kroenke K, Williams JBW. Patient Health Questionnaire Primary Care Study Group. Validation and utility of a self-report version of PRIME-MD: The PHQ primary care study. JAMA. 1999.282(18), 1737-1744. 10.1001/jama.282.18.1737

28. Yesavage JA, Brink TL, Rose TL, et al. Development and validation of a Geriatric Depression Screening Scale: A preliminary report. Journal of Psychiatric Research. 1982; 17(1), 37–49. 10.1016/0022-3956(82)90033-4

29. UDDIN, Md Zia, et al. Deep learning for prediction of depressive symptoms in a large textual dataset. Neural Computing and Applications, 2022, 34.1: 721–744.

30. Guohou S, Lina Z, Dongsong Z, What reveals about depression level? The role of multimodal features at the level of interview questions. Information & Management. 2020;57, Issue7

31. Amanat, A., Rizwan, M., Javed, A. R., Abdelhaq, M., Alsaqour, R., Pandya, S., & Uddin, M. (2022). Deep learning for depression detection from textual data. Electronics, 11(5), 676.

